# The COVID-19 Pandemic Predominantly Hits Poor Neighborhoods, or does it? Evidence from Germany

**DOI:** 10.1101/2020.05.18.20105395

**Authors:** Thomas Plümper, Eric Neumayer

## Abstract

**Background:** Reports from the UK and the USA suggest that COVID-19 predominantly affects poorer individuals and neighbourhoods. This article paints a more complex picture by distinguishing between a first and second phase of the pandemic. The initial spread of infections and its correlation with socioeconomic factors largely depends on how the virus first entered a country. The second phase of the pandemic begins when individuals start taking precautionary measures and governments implement lockdowns. In this phase the further spread of the virus depends on the ability of individuals to socially distance themselves, which is to some extent socially stratified.

**Methods:** We analyze the geographical distribution of known cases per capita across 401 local districts in Germany, once for infections in the initial phase and for new infections during the second phase.

**Results:** In Germany, the virus first entered via individuals returning from skiing in the Alps and other international travel. In this first phase we find a positive association between the wealth of a district and infection rates and a negative association with indicators of social deprivation. During the second phase, richer districts and districts with a higher share of university-educated employees record fewer new infections, whilst the initial safety advantage of more socially deprived districts disappears.

**Conclusion:** The social stratification of Covid-19 changes substantively across the two phases of the pandemic in Germany. Only in the second phase does socio-economic advantage turn into a safety advantage. Thus, suggestions that the pandemic predominantly hits the poor needs to be qualified.

## Introduction

It is a recurrent theme in the public health literature that during a pandemic, low-income populations will be infected with a higher probability (Bouye et al. 2009; Bernardi 2020; van Dorn et al. 2020). In line with this established argument, recent media reports from the United Kingdom (Toynbee 2020), data from the US (Han et al. 2020) and an academic study of Covid-19 mortality in New York City (Harris 2020) suggest that the poor are more likely to get infected with Sars-CoV-2 and develop Covid-19 and that poor neighborhoods will bear the brunt of the pandemic. In other countries, however, Covid-19 is perceived to be a ‘rich man’s disease’. In many developing countries, the coronavirus was imported via business travelers from China, students from Europe, or by tourists (Bengali et al. 2020), and in European countries, a good part of the spread can be traced back to ski tourism in the Alps (Correa-Martínez et al. 2020).The argument that Covid-19 affects and kills predominantly the poor and socially deprived cannot be generalized to all countries and the link between socio-economic factors and who bears the major burden of the pandemic is more complex. Our argument rests on a distinction between two phases of the pandemic. In phase 1 the first importation of the virus from abroad and its subsequent initial spread will depend to some extent on pure chance and its correlation with socio-economic factors in a country is strongly influenced by the channels through which the virus enters a country. This initial phase is characterized by widespread ignorance first and neglect later on. The existence of the virus may be known – European citizens had seen footage from China and Italy – but the ability to test for a Sars-CoV-2 infection remains underdeveloped (Gross et al. 2020) and very few people had adjusted their behavior and governments had not yet implemented social distancing measures.

In Germany, the virus happened to be spread initially via individuals returning from ski holidays in the Alps and, to a much lesser extent, through business and other travelers from China, Italy and other hotspots, which meant that the majority of infected people in the beginning were relatively young and well-off (Correa-Martínez 2020; Steffens 2020). In Italy, by contrast, the virus was initially spread mainly through hospitals, which made the average infected Italian much older than the average infected German. Once the virus had reached Germany, the subsequent spread of infections was facilitated by super-spreader social events such as a carnival session in Gangelt, a small town in the district of Heinsberg on the border to the Netherlands, a beer festival in the small city of Mitterteich, district of Tirschenreuth, and a wine event in Bretzfeld, Hohenlohekreise. These super-spreader events create local cluster effects if the social event is mainly attended by locals. In fact, even two months after the above events took place, these were still the districts with the highest number of known infections per 100,000 citizens in Germany.

Map 1 displays known Sars-CoV-2 cases, normalized by population, in German districts on 13 April. Even at a first glance we see that the rate of infection declines from South to North and from West to East. Even within the Western part of Germany, regions in which a greater share of the population is Catholic also have a higher incidence, which may be correlated to spreader events such as carnival that is much more popular in predominantly Catholic regions. The North-South divide appears to be stronger than the East-West divide. This may be down in part to the greater ease by which Southern Germans can reach by car what turned out to be virus hotspots in ski resorts in Northern Italy and Austria.

**Map. 1.**
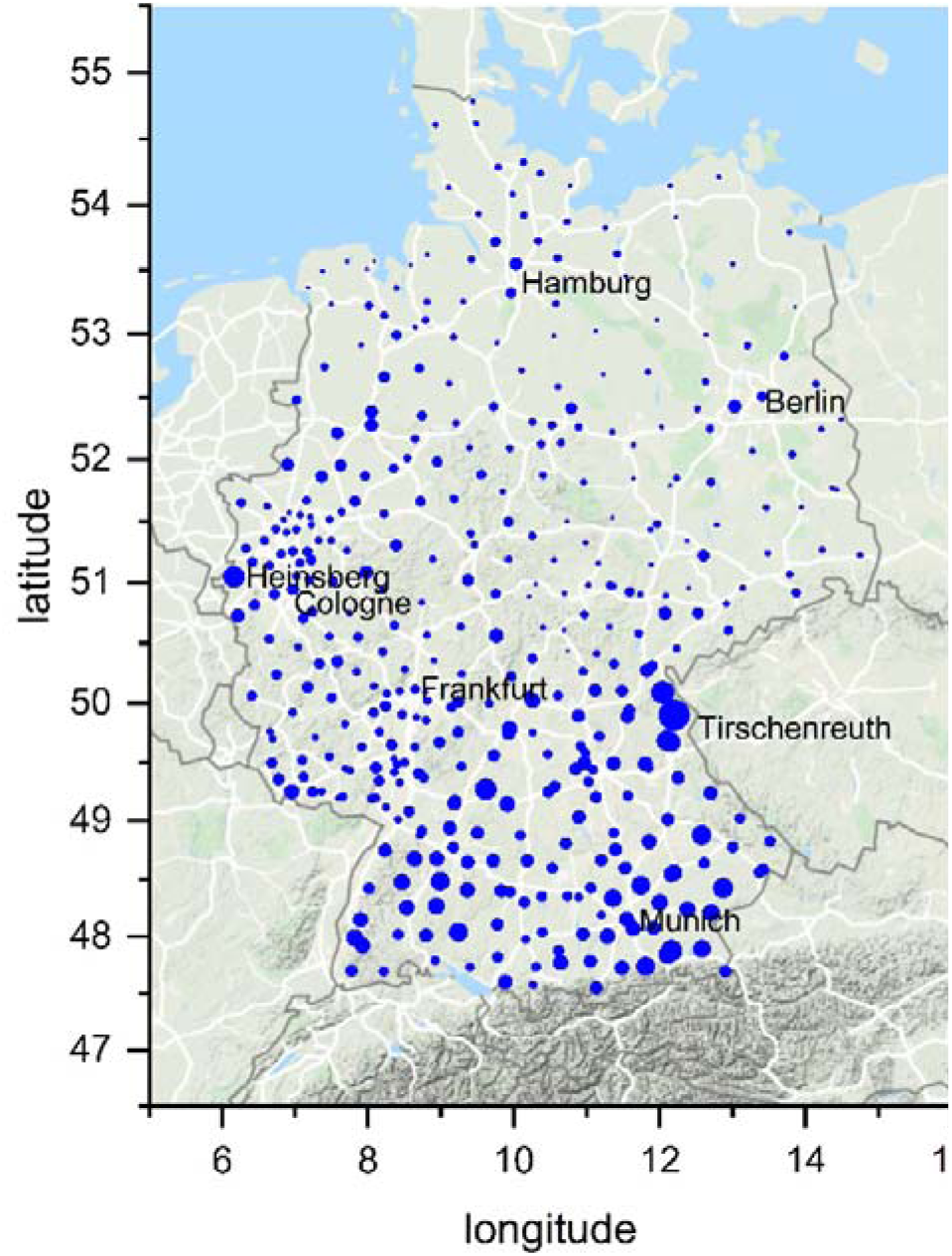
Distribution of Known Sars-CoV-2 Infections Per Capita, April 13

Once the existence and dangers of the pandemic have become public knowledge, people and governments implement precautionary measures and the spread of the virus slows down (Maier and Brockmann 2020; Schmitt 2020). At the same time, the geographical pattern of infections slowly changes. For a virus to spread, social interaction between an infected and an uninfected person is required. Since the number of new infections remains strongly influenced by the number of active infections in a district, the pattern that has evolved during phase 1, will not disappear quickly. Thus, hotspots remain hotspots for some time.

But not forever. Map 2 shows the distribution of new positive Sars-Cov-2 tests between 13 April and 17 May. Whilst the North-South and East-West divides are still present, clearly the distribution of new infections already starts to look different from the distribution of cases on 13 April.

**Map. 2.**
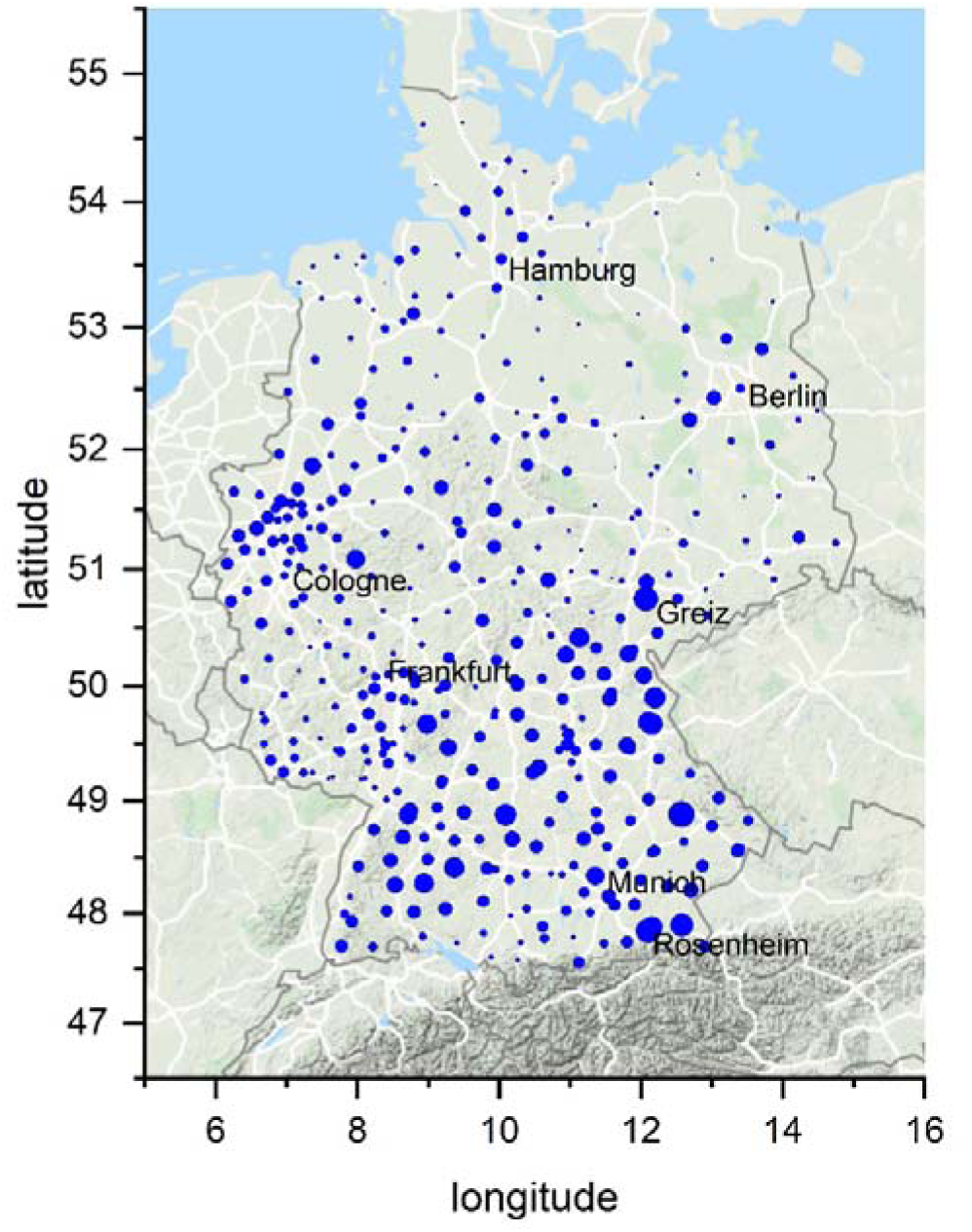
Distribution of New Known Sars-CoV-2 Infections Per Capita, 13 April to 17 May

In phase 2 of the pandemic and despite the enduring and strong path-dependency, which map 2 visualizes, the correlation of the spread of the infection with socioeconomic factors changes. In the second phase, individuals reduce social interactions, governments implement lockdown measures and recommend social distancing especially to people perceived as vulnerable. During the second phase of the spread of the virus, the ability of individuals to reduce social interactions becomes the decisive factor. Lockdown measures do not affect all people in the same way (Glover et al. 2020).

The ability to reduce social interactions and to ‘stay home’ is not distributed evenly in a society (Almagro and Orane-Hutchinson 2020a, 2020b). The spread of the virus in phase 2 is shaped by the extent to which individuals manage to reduce their social contacts. In general, white collar activities can be moved to a home office, while other workers still need to commute to their workplace and work if their employer does not lock down the workplace. Poorer people find social distancing more challenging than richer people, having less access to resources to shield them from the economically damaging effect of the lockdown. The further spread of the virus becomes increasingly socially regressive. Regardless of how and where the virus had spread first in the initial phase of the pandemic, in phase 2 the virus is likely to become a poor man’s disease.

To test these predictions, we study the social stratification and the dynamics of the coronavirus pandemic in Germany. Specifically, we analyze the geographical distribution of known coronavirus cases across 401 local districts in Germany. We find that, firstly, the initial risk of becoming infected during phase 1 was higher in richer districts and was lower in more socially deprived districts. Secondly, we also provide evidence of the socially regressive nature of new infections once phase 2 has begun. Richer districts and districts with a higher percentage of the workforce who are university educated now see lower new infections, whereas the initial safety advantage of more socially deprived districts vanishes.

## Methods

The transition from phase 1 to phase 2 is a smooth process rather than a hart cut, as this depends on when people start consciously changing their behavior and some do so earlier than others. Still, a definite break comes with the lockdown. The first German states to go into lockdown were Bavaria and the Saarland. Their curfew begun on 21 March; one day later the whole of Germany followed. Hartl et al. (2020: 26) find that “confirmed Covid-19 cases in Germany grew at a daily rate of 26.7% until 19 March. From March 20 onwards, the growth rate drops by half to 13.8%, which is in line with the lagged impact of the policies implemented by the German administration on 13 March and implies a doubling of confirmed cases every 5.35 days. Before 20 March, cases doubled every 2.93 days.”

Ideally, therefore, we would test our first prediction with data on cases from late March or early April, since it takes roughly a week from the implementation to the effectiveness of policy measures on infection rates. Unfortunately, the first date at which we were able to capture the full distribution of confirmed infections across all German districts is 13 April, with data sourced from the website of the Robert Koch Institute. Whilst clearly introducing measurement error as overlapping with the second phase of the pandemic, the strong path dependency of any pandemic means that the stock of infections on 13 April will be sufficiently strongly correlated with the stock of infections around 30 March, which would have been the ideal date.

To study our second prediction, we take as our second dependent variable new infections that happened in the second period between 13 April and 17 May. These new infections all occurred after people had time to adjust to the by now fully known risks and the lockdown had been imposed. We divide the initial stock of infections and the new infections thereafter by population size in a district in 10,000 people. Consequently, the dependent variables in our regressions represent cases and new infections per capita. We estimate our regression models with ordinary least squares and robust standard errors.

As our measure of wealth of a district we include the average income subject to income tax in thousands of Euro. We also control for the share of the workforce that is university-educated. This variable is a proxy for the share of the population that can move work from a home office. To measure social deprivation we include two variables, namely the percentage share of the population in a district that receives social welfare benefits as well as the unemployment rate. For brevity, we only show results for the first variable but results are qualitatively similar if we include the unemployment rate instead. Average taxable income is highly negatively correlated with the population share of social welfare benefits and the unemployment rate at r = −0.46 and r = −0.58, which is why we include average taxable income and the population share of social welfare benefit recipients only in separate regressions.

As two, admittedly crude, measures to account for the way in which the virus first entered Germany and spread initially we include the latitude location of a district and the share of its population that is Catholic. The former accounts for the ease by which residents could drive to the Alps for ski tourism, whilst the latter account for the greater popularity of carnival as potential super-spreader events in predominantly Catholic districts. A further way in which we account for the path dependency of the pandemic is by including the stock of cases on 13 April into the regressions with the new infections between 13 April and 17 May as dependent variable. The correlation between April 13 infections and new infections on May 17 is approximately 0.59. The variable can function as a proxy for the risk of new infections stemming from existing infectious cases in the district.

In addition, we include dummy variables for the nature of habitation of a district, namely whether it is predominantly urban, and for the extreme remoteness of a district. The virus should spread more easily in more densely populated urban habitats (Zelner et al. 2012; Ameh 2020) and while extreme remoteness is often seen as a costly locational disadvantage (Redding and Sturm 2008; Dasgupta et al. 2012), it partly protects the local population from infections as there will be less exchange with people from the outside. All data for the explanatory variables are sourced from regional databases of the German statistical offices.

## Results

Table 1 reports results for average taxable income as the central socio-economic explanatory variables, table 2 does the same for the population share of social welfare benefits. Both sets of regressions contain the university-educated share of the workforce. Average taxable income is positively associated with a higher share of cases on 13 April in a district. A district that is richer on average by 10,000 Euros is estimated to have 5.6 additional cases per 10,000 people. The mean of cases per 10,000 people on 13 April is 14.9 with a standard deviation of 12.3. By stark contrast, average taxable income is negatively associated with a higher share of new infections after 13 April. A district that is richer on average by 10,000 Euros is estimated to have 1.6 fewer new infections per 10,000 people. The mean of new infections per 10,000 people is 6.2 with a standard deviation of 5.7. Richer districts were initially more affected but manage to have fewer new infections than poorer districts in the second phase of the pandemic. The same is true for districts with a higher share of university-educated workforce. A ten percentage points increase in this share is associated with 1.4 fewer infections per 10,000 people.^1^

**Table 1.** The evolving effect of average taxable income.

| Dependent variable: | (1)<br>Cases per capita 13<br>April | (2)<br>New infections p.c. to<br>17 May |
| --- | --- | --- |
| Cases per capita on 13 April |  | 0.263***<br>(0.0451) |
| Taxable income per capita | 0.556***<br>(0.165) | -0.160**<br>(0.0776) |
| % workforce university-educated | -0.0801<br>(0.105) | -0.137**<br>(0.0576) |
| Urbanity dummy | -0.810<br>(1.140) | 1.010*<br>(0.580) |
| Extreme periphery dummy | -1.362<br>(1.142) | -1.752***<br>(0.503) |
| Latitude | -1.898***<br>(0.369) | -0.442**<br>(0.197) |
| Population share catholic | 0.135***<br>(0.0446) | -0.0143<br>(0.0150) |
| Constant | 97.89***<br>(19.54) | 28.99***<br>(10.88) |
| Observations | 401 | 401 |
| R-squared | 0.337 | 0.388 |

Note: Robust Standard errors in parentheses. \*\*\* p<0.01, \*\* p<0.05, \* p<0.1

**Table 2.** The evolving effect of social welfare benefits per capita.

| Dependent variable: | (3)<br>Cases per capita 13<br>April | (4)<br>New infections p.c. to<br>17 May |
| --- | --- | --- |
| Cases per capita on 13 April |  | 0.262***<br>(0.0435) |
| Social welfare benefit claimants p.c. | -0.480***<br>(0.124) | 0.134*<br>(0.0800) |
| % workforce university-educated | 0.0978<br>(0.103) | -0.187***<br>(0.0582) |
| Urbanity dummy | 0.525<br>(1.184) | 0.633<br>(0.611) |
| Extreme periphery dummy | -1.966*<br>(1.132) | -1.579***<br>(0.500) |
| Latitude | -1.656***<br>(0.429) | -0.511**<br>(0.201) |
| Population share catholic | 0.147***<br>(0.0414) | -0.0175<br>(0.0158) |
| Constant | 96.74***<br>(22.60) | 29.29***<br>(10.69) |
| Observations | 401 | 401 |
| R-squared | 0.335 | 0.375 |

Note: Robust Standard errors in parentheses. \*\*\* p<0.01, \*\* p<0.05, \* p<0.1

The opposite pattern emerges for the population share of social welfare benefits as the central explanatory variable - see table 2. Districts with a larger population share of social welfare benefit recipients initially had a safety advantage as they report lower cases on 13 April. One fewer case per 10,000 people is associated with a two percentage points increase in the population share of social welfare benefit claimants among the district population. This statistically significant negative association disappears for new infections in model 4 and actually turns positive, if only marginally statistically significant. More socially deprived districts lose their initial safety advantage in the second phase of the pandemic.

We find that latitude maintains its negative association with the initial stock of cases on 13 April and with new infections thereafter. The catholic population share has a statistically significant positive association with initial cases but not with new infections. The urbanity of a district is not statistically significantly associated with the initial stock of cases on 13 April and only marginally so with new infections in model 2. The 18 districts that are considered to be located in the extreme periphery of Germany see fewer new infections but the negative association for cases on 13 April is only marginally statistically significant in model 3.

## Discussion

The COVID-19 pandemic does not simply predominantly affect poorer and socially deprived neighborhoods more than richer and less deprived ones. Evidence from the US and the UK suggests that such an outcome is possible, but as the evidence from Germany presented in this article demonstrates, one cannot generalize to other country settings. In Germany richer districts were more affected and socially deprived neighborhoods were less affected in the first phase of the pandemic mainly because ski tourism is expensive and because the districts geographically closer to the Alps are also relatively wealthy and have little social deprivation.

This pattern started to change in the second phase of the pandemic following the lockdown. The safety advantage that socio-economically disadvantaged districts had in the initial phase disappears in the second phase. Controlling for the strong path dependence in infections data, during the lockdown phase of the pandemic people in richer districts as well as districts with a higher share of university educated employees find it easier to protect themselves. In Germany at least, only in the second phase of the pandemic does socioeconomic disadvantage go hand in hand with a safety disadvantage.

## Data Availability

Data used in this article and do files that allow to replicate the reported results can be obtained from the authors.

## Footnotes

1 Our results are substantively similar to those reported in tables 1 and 2 if we include fixed effects for the 16 states that make up the Federal Republic, except that the positive association of average taxable income with the initial stock of cases becomes statistically insignificant.

